# Assessment of adherence to drug regimen, its potential factors and quality of life among patients who have undergone myocardial revascularization

**DOI:** 10.1101/2025.07.08.25331154

**Authors:** Bhanumati Dutta, Pares Bandyopadhyay, Uma Adhikari

**Affiliations:** The West Bengal University of Health Sciences; CTVS Dept., NRS Medical College and Hospital, Kolkata, W.B.; College of Nursing, NRS Medical College and Hospital, Kolkata, W.B.

**Keywords:** Coronary artery Disease, Adherence to drug regimen, myocardial Revascularisation Percutaneous transluminal coronary Angioplasty, Coronary Artery Bypass surgery, Quality of Life

## Abstract

**Introduction:** Quality of life is the ultimate outcome for a patient who has undergone myocardial revascularization. Among several influencing factors, adherence to drug regimen enhances its level by preventing restenosis of stent or graft and restricting the progress of native CAD. Nurse’s role is to assist patient with myocardial revascularization for maintaining high quality of life by exploring the associated issues in a greater extent.

**Objectives:** The objectives of the study were to assess the level of adherence to drug regimen, to explore potential factors of adherence to drug regimen and to assess quality of life among patients who have undergone myocardial revascularization.

**Methodology:** A quantitative research approach and prospective longitudinal cohort study design was adopted to collect data from three hundred and fifty (355) patients who have undergone either PTCA or CABG, selected by nonprobability convenience sampling technique. ARMS, SF-36 short form and an interview schedule were used to collect data from the subject at three months’ interval till nine (9) months after revascularisation.

**Result:** The study result shows that majority (82%) of the patients with myocardial revascularisation were adherent to drug regimen at initial month after revascularisation but reduced to 52% after nine months(p<0.01). The percentage of adherence with medication was significantly reduced to 74% after 3 months, 75% after 6 months, and 59 % after 9 months among patients with CABG whereas percentage of adherence among patient with PTCA was also reduced to 66% after 3 months, 63% after 6 months and 44 % after 9 months (p<0.01)

Age and educational status were independent predictive factors for adherence to drug regimen among patient with PTCA.

The potential associated factors related to adherence to drug regimen was no of drugs unavailable at Govt. medicine counter.

The two main domains, of quality of life as Physical component summary and Mental component summary of patient with myocardial revascularisation were improved from one month to 9 months after intervention (p<0.001)

Physical component summary of Quality of life had a negative relationship with adherence to drug regimen at 3 months, 6 months and 9 months’ time period whereas mental component summary of Quality of life had a significant negative relationship with adherence to drug regimen at 6 months’ time period.

**Conclusion:** Adherence to drug regimen was decreased over time but Quality of life was improved among patients with myocardial revascularisation. There was a positive relationship with adherence to drug regimen with PCS domain of quality of life among patients with myocardial revascularisation. Age and educational status were independent predictive or potential factors for adherence to drug regimen among patient with PTCA.

## Background of the study

Coronary artery disease(CAD) is continuing as a leading cause of mortality and morbidity worldwide as more than half a billion people all over the world continue to be affected by cardiovascular diseases(CVD), which was accounted for 20.5 million deaths in the year [1]. Based on the report of the World Health Organization, India accounts for one-fifth of deaths due to cardiovascular disease worldwide, especially in younger population due to increasing incidence of modifiable and non-modifiable risk factors. [2,3]

The existing treatment of CAD is myocardial revascularisation either in the form of medical, surgical or a combination of both depending on clinical presentation, extent and severity of the disease. Myocardial revascularisation is the procedure to restore blood supply to ischemic myocardium of patients with CAD or Acute Coronary Syndrome(ACS) in an effort to limit ongoing damage, reduce ventricular irritability, and improve short-term and long-term outcomes [11] Myocardial revascularisation in the form of Coronary Artery Bypass surgery (CABG), Percutaneous coronary intervention (PCI) and emerging different novel hybrid procedures like stem cells therapy, nanotechnology, robotic surgery and 3-D printing, are showing promising effects in managing CAD in both developed and developing countries.[6]

The recent data shows that CABG and PCI remain the most common management of CAD in developing countries like India, China, Pakistan etc. Annually almost 8 lakhs patients in USA, 9 lakhs in China, 2.5 lakhs patients in Japan and over 1.2 million patients with CAD in Europe undergo Myocardial revascularisation either in the form of CABG or PCI (5)

But the success of the procedure or intervention depends on secondary prevention by medication therapy as patients sustain with a risk of subsequent ischemic events, resulting from progression of native CAD, development of atherosclerosis in venous graft and stent thrombosis. [6,7]

The combination of aspirin and clopidogrel as dual antiplatelet therapy and lipid-lowering therapies [7] continue to be the mainstay of secondary prevention which will help to maintain long-term graft patency and assist patients to obtain the highest level of physical health and quality of life following CABG and PCI. Antihypertensive drugs-angiotensin converting enzyme inhibitors (ACEI), angiotensin II receptor blockers [9] glucose-controlling agents [8], smoking cessation, weight loss, [7] use of beta-blocker [9] and cardiac rehabilitation are considered as essential for long time survival and to limit complications of patients who have undergone CABG or PCI [9]

A single centre cohort observational study conducted on Medication adherence on three reimbursed drugs as ACEIs (ramipril, perindopril), P2Y12 receptor inhibitors (clopidogrel) and statins (atorvastatin, simvastatin, rosuvastatin). and its determinants among post-MI patients treated with primary coronary intervention (PCI) showed that in 1-year follow-up, for all three drug classes level of adherence was 64 ± 25%, with 67 ± 32% for ACEIs, 62 ± 34% for P2Y12 receptor inhibitor and 64 ± 32% for statins. A gradual decline in adherence was found from 65% ± 26% in the first quarter of follow-up to 51% ± 34% in the last quarter of follow-up (p < 0.00001). Sufficient adherence for all drugs classes was found only in 29% of patients throughout the whole follow-up period (44% for ACEI, 36% for P2Y12 receptor inhibitor and 41% for statins).The independent predictors of drug adherence were age, prior CABG, level of education, place of residence, economic status and marital status Whereas patients > 65 years and having a prior history of CABG more often had an insufficient adherence to drugs, married and hypertensive patients, city inhabitants and patients with higher education showed to have a sufficient drug adherence. [34]

The result of an another study conducted on Medication Adherence and its Related Factors in Patients Undergoing Coronary Artery Angioplasty indicated that 28% patients did not adhere to their medication and medication adherence was associated with the patient’s spouse’s education (P=0.04), the family history of coronary artery disease (P=0.03), the history of hypertension (P=0.09) and significantly related only among the male patients (P=0.025).The independent predictors of the medication adherence were only the spouse’s education and the family history of CAD.[33]

Several studies reported high levels of nonadherence to drug regimen among patients with CAD, in the range of 33% to 50%, which is associated with10-40% relative increase in the risk of rehospitalisation and a 50% to 80% relative increase in mortality. [10]

But the adherence to medication encompasses a complex interplay of several factors involving sociodemographic, person related, health care delivery system related, health care provider and medicine consumption related factors [56, 57,58]

The rate of non-adherence to at least one Secondary prevention medicine(SPM) among patients with CAD was 43.5% (n=219), but 53.3% of reported non-adherence was to only one SPM. The related factors were, forgetfulness (84.9%; n=186), worry that medicines will do more harm than good (33.8%; n=74), feeling hassled about medicines taking (18.7%; n=41), feeling worse when taking medicines (14.2%; n=31) and not being convinced of the benefit of medicines (9.1%; n=20). [9]

But important outcome measure after any intervention is the patients’ subjective appraisal of the intervention. [59]

The World Health Organization (WHO) defined health as a complete physical, mental and social well-being not merely the absence of disease. So the quality of life is the most effective patient oriented methodology or criterion to measure therapeutic achievements than different physician centred methodology. Good QOL implies the person’s ability to function normally on a daily basis and to be satisfied with the participation in daily activities which includes preserved physical mobility, independence, sufficient energy for self-care activities, social contacts, emotional stability, absence of pain or other symptoms of discomfort and adequate sleep and rest. [59]

A study conducted on quality of life in patients of different age groups before and after Coronary Artery By-Pass Surgery showed the enhanced QOL six months to 1 year after CABG surgery in the majority of patients regardless of age [38]

But another study showed that quality of life after CABG is not improved in all domains, and some patients even experience poorer health-related quality of life (HRQOL) after the surgery [6,7]

A single centre observational study conducted on Determinants of non-recovery in physical health-related quality of life one year after cardiac surgery among 1920 patients shows that in comparison to the preoperative status, 176 (21.9%) patients one year after cardiac surgery did not display an improvement in the SF-36 physical domain score. For isolated CABG, 23.2% of patients did not display any improvement in the physical domain score. []

A study conducted among patients who have undergone PTCA showed that the mean score of the patients’ quality of life in three domains were 67.16 ± 12.04 (social functioning), 64.33 ± 8.73 (physical functioning), 65.45 ± 9.31 (emotional functioning), and 126.6 ± 15.99 in total quality of life score. In multivariate analysis it is revealed that age (*P* = 0.0001), the number of diseased vessels (*P* = 0.0001), and the number of comorbidities (*P* < 0.05) were the most important factors associated with the quality of life in patients undergoing Percutaneous coronary angioplasty.

Several factors might be associated with quality of life in patients undergoing coronary angioplasty A retrospective cohort study by Hasandokht T, Salari A. on Medication adherence and quality of life in coronary artery bypass grafting patients revealed that Physical and mental components of QOL were negatively associated with medication (B:-0.18, p:0.04; B:-0.29, p:0.02, respectively) and follow-up visit observance (B:-0.3, p:0.01; B:-0.3, p: 0.01, respectively).[19]

So different studies highlighted that quality of life after myocardial revascularisation had association with adherence to medication and several factors may change the level of adherence.

### Need of the study

Coronary heart disease (CHD) is now the leading cause of death worldwide. According to World heart report 2023, An estimated 3.8 million men and 3.4 million women die each year from CHD. In Europe CHD accounts for an estimated 1.95 million deaths each year. CHD is the most common cause of deaths in the UK. An estimated 1 in 5 men and 1 in 6 women die from the disease each year. In 2003 CHD caused around 114,000 deaths in the UK.CHD is responsible for 110,000 deaths in England each year [59]

The results of Global Burden of Disease study stated that age-standardized CVD death rate in India was 272 per 100000 populations which is much higher than that of global average death rate of 235[1].

The American Heart Association (AHA) and the World Health Organization recognize the key role that nurses, and other team members play in supporting the goal to reduce death and disability from CVD by 25% in 2025. For more than 4 decades, nurses and advanced practice nurses have taken on key roles in managing single and multiple risk factors, including hypertension, smoking, lipids, and diabetes; the sequelae of chronic conditions, such as coronary artery disease and heart failure, through specialized clinics; and programs in primary care, worksites, and cardiac rehabilitation [22]

By taking on a primary role as team leaders in providing case management, nurses have proven their capability to not only reduce CVD risk factors, but to also adhere to treatment guidelines and protocols, decrease hospitalization, and reduce morbidity and mortality in those with established disease. Such programs demonstrating improved outcomes and cost effectiveness have been noted in both developing and developed countries [22]

Therefore more studies need to be conducted to find out the level of adherence to drug regimen what are the potential factors and how it is related to the quality of life of the patient. Majority of the patients in middle and low income group receive the health facilities from Government hospital and unlike the follow up procedures conducted by community health care provider in USA,U.K. and other developed countries,regular follow up in Indian scenario was done at hospitals only. But very less studies have been conducted to highlight this issue in developing countries like India, Pakistan, Bangladesh etc. as well as in West Bengal, covering both the groups of patients who have undergone PTCA or CABG. So the researcher felt the need to conduct the present study at the Outpatient Department(OPD) of Government hospital to find the real situation which will help to formulate new policy for enhancing better quality of life after Myocardial revascularisation.

## Title of the study

Assessment of Adherence to drug Regimen, its potential factors and quality of life among patients who have undergone myocardial revascularization attending cardiac OPD of NRS Medical College and Hospital of Kolkata, West Bengal

## Objectives

Primary objectives

- To assess the level of adherence to drug regimen among patients who have undergone myocardial revascularisation
- To find potential factors related to adherence to drug regimen among patients who have undergone myocardial revascularisation
- To assess quality of life of patient who have undergone myocardial revascularization. Secondary objectives
- To find association between level of adherence to drug regimen with potential factors
- To find association between level of adherence to drug regimen among patients who have undergone myocardial revascularization with selected demographic variables
- To find association between quality of life of patients who have undergone myocardial revascularization with selected demographic variables
- To find relationship between level of adherence with Quality of life

## Research variables

There were three variables in the present study

- Socio demographic variables including potential factors
- Adherence to drug regimen
- Quality of life

## Hypotheses

All the hypotheses would be tested at 0.05 level of significance

H_1_ There is a significant difference in level of adherence to drug regimen between initial month, after three months, after six months and nine months among the patient who have undergone Myocardial revascularisation (CABG or PTCA)

H_2_ There is a significant association between level of adherence to drug regimen with potential factors such as sociodemographic, condition related, therapy related, person related, provider and health care delivery system related factors.

H_3_ There is significant correlation between level of adherence to drug regimen with PCS and MCS of Quality of life among patients who have undergone Myocardial revascularisation.

## Operational Definitions

### Adherence to drug Regimen

In the present study adherence to drug regimen is the degree to which a patient follows medication schedule. It is referred as prescription to be obtained and drugs to be taken as prescribed – in terms of dose, dosing interval, duration of treatment and any special instructions in taking medicine and refilling of medicine, which is the main issue of developing country. So Adherence to refills and medications scale (ARMS) tool would be used to assess the level of medication adherence.

### Potential factors

In the present study potential factors of medication adherence meant which have effect on adherence to drug regimen and composed of five significant factors.

**Socio-demographic factors included** demographic, social and economic variables **condition related factors** as illness representation with comorbid conditions, **therapy related factors** as medicine characteristics and consumption of medicine, **Patient or Person related**

**factors** as person providing medications,cost of purchasing unavailable medicine from Govt. medicine counter and knowledge about drugs and **Provider and health care delivery system related factors** included no of medicines unavailable in Govt. medicine counter and Information provided by counter/Doctor/Pharmacist regarding time and effects of medicine and which would be assessed by a self-developed Interview schedule. [68]

### Quality of life

It is a broad multidimensional concept that usually includes subjective evaluations of both positive and negative aspects of life as physical functioning, psychological wellbeing, social and role functioning and health perceptions In the present study it would be assessed by SF-36 Health related quality of life tool. [67]

### Patient undergone Myocardial Revascularisation

The patients who have undergone procedures that restore blood supply to the diseased myocardium which can be achieved through various methods, including surgery, bypass procedures, or minimally invasive techniques like angioplasty and stenting myocardium by bypassing or removing the blockage of affected coronary arteries and attending cardiology or Cardiovascular and Thoracic surgery OPD for follow up. [61]

## Literature Review

The research and non-research literature were reviewed and organized under the following headings

- Prevalence of myocardial Revascularization
- Adherence to drug regimen among patient who have undergone CABG or PTCA
- Potential factors related to adherence to drug regimen
- Factors associated with non-adherence to cardiovascular medicines.
- Quality of life of patients who have undergone Myocardial revascularization-CABG or PTCA

## Research Methodology

**Fig 2.**
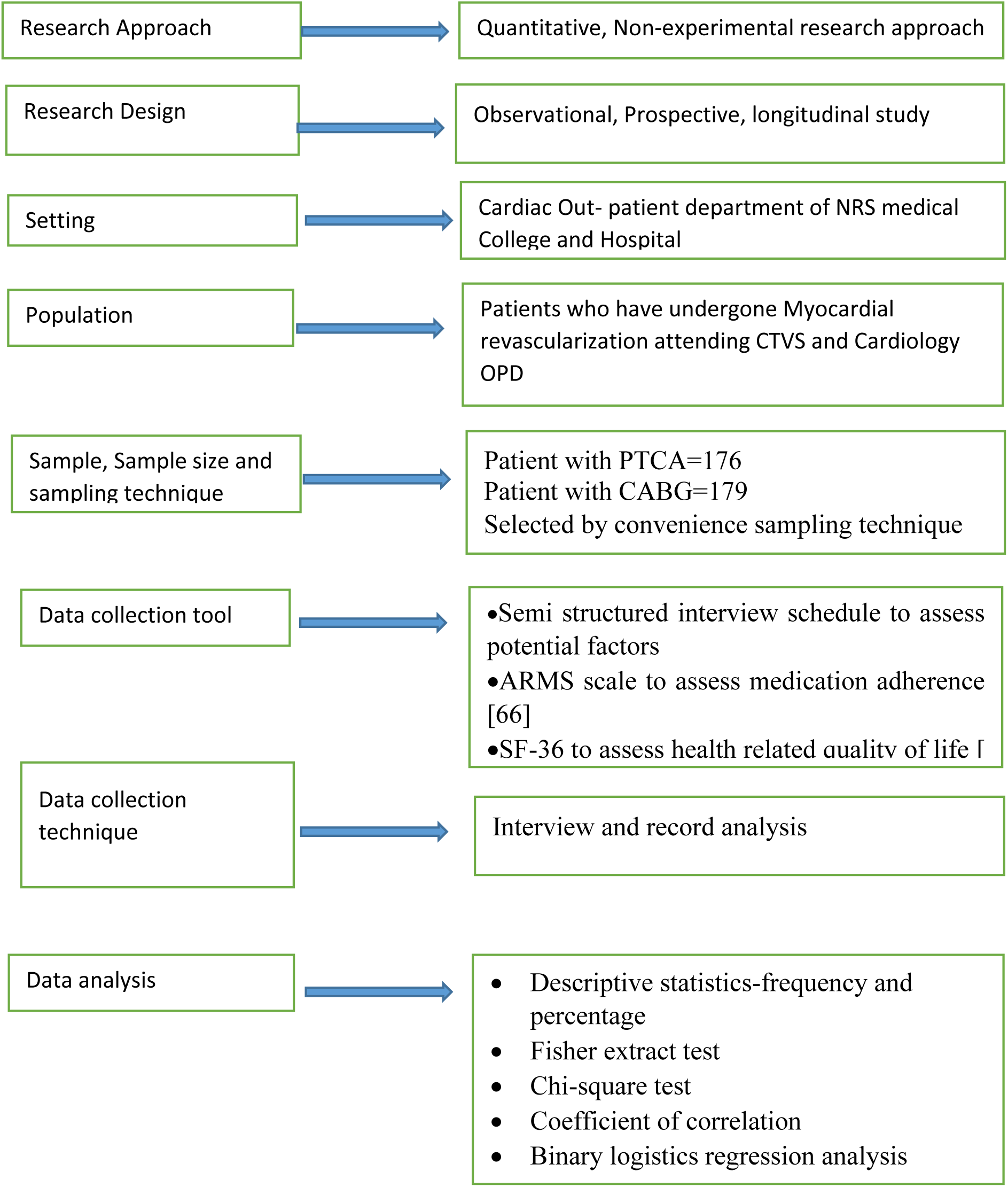
Schematic diagram of research design.

### Sample size

Formula used to calculate sample size for descriptive study

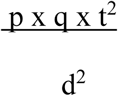

p=prevalence of adherence to cardiovascular medication 0.65 []

q= 1-p

t= critical value at 0.05 level of significance

d=level of significance

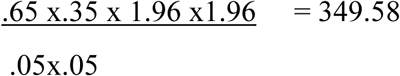

Sample mortality 15%=52.43

So total sample size was 349.58+ 52.43 = 402.017=403

#### Recruitment of the subjects

The patients meeting inclusion and exclusion criteria were grouped in a cohort. The purpose of the study was explained by the investigator. The willing subjects were recruited. There were major two separate cohorts. One cohort was made by the subjects who have undergone CABG and another cohort was made by the subjects who have undergone PTCA.

**Fig 3.**
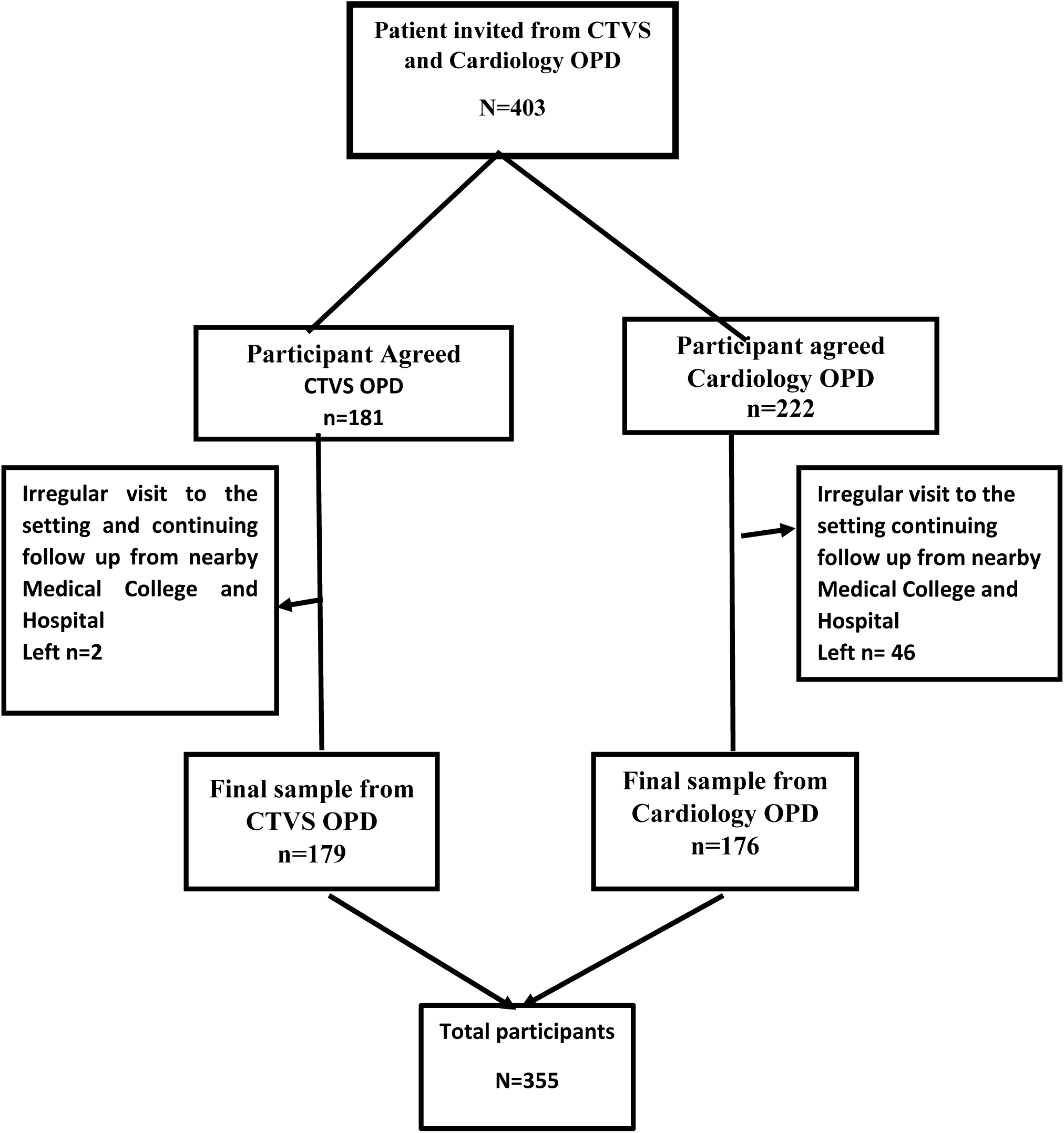
Subject recruitment process.

## Translation of the tools

The three validated tools were translated from English to Bengali version and retranslated to English version to establish linguistic validity. The translation of tools was done following the proposed guidelines by Beaton DE, Bombardier C, Guillemin F, Ferraz MB. [65].

## Pretesting

The three validated and translated tools were pretested from 12^th^ to 16^th^ November 2019 among 10 patients who have undergone either CABG or PTCA attended cardiac OPD at NRS Medical College and Hospital. The total time taken to complete the tools by interview technique and record analysis was 40 minutes. All the items were reported clear to the subjects with no ambiguity.

## Reliability of tools

The reliability was computed for knowledge questionnaire of part D of Tool I. The coefficient of correlation (α) was 0.84, computed by Cronbach’s alpha method to establish the internal consistency of seven (7) items of Knowledge questionnaire. Hence the knowledge questionnaire was found to be reliable.

The two tools ARMS and SF 36 short form of English version were standardized scales with established reliability. The reliability of Bengali version of both the tools were established for internal consistency. The coefficient of correlation (α), computed by Cronbach’s alpha method was 0.82 and 0.89 respectively. Hence both the Bengali version of tools seemed to be reliable.

## Pilot Study

The pilot study was conducted in the month of January, 2020 to April 2020 at NRS Medical College and Hospital. Twenty (20) patients who had come for follow up after CABG and Twenty (20) patients who had come for follow up after PTCA were included in the cohort. The non-probability convenience sampling technique was used to select the subjects from the CTVS and Cardiology OPD.

Pilot study result showed that Adherence to drug regimen among patients with CABG and PTCA were 84% and 86% respectively. There was no association between adherence to drug regimen with potential factors for whole group and also in individual group of patient with CABG or PTCA. The PCS and MCS quality of life were improved among whole and both the group after surgery or intervention. But no relation was found in PCS and MCS of quality of life for whole and individual group with adherence to drug regimen.

## Ethical Consideration

The ethical permission was obtained from –

- Institutional Ethics committee of NRS Medical College and Hospital
- Written informed consent from individual subjects, after explaining the information guide sheets in Bengali and Hindi as per language spoken.
- Anonymity and confidentiality were maintained throughout the study.

## Data collection procedure for final study

The main study data collection was commenced from January 2021 and completed in the month of April 2023 at NRS Medical College and Hospital. The patients who had come for follow up after CABG and after PTCA was included in two major cohorts. The non-probability convenience sampling technique was used to select the subjects from the CTVS and Cardiology OPD.The researcher introduced herself and purpose of the study was explained. After the doctor’s visit they were interviewed in a separate small room at CTVS department in the OPD no 5 and also at the corner of the cardiology OPD room. The written informed consent was obtained from individual subject. The three (3) tools were administered by interview technique and clinical records were analyzed from discharge paper, OPD sheet and medicine slip written by the Physician. The total time taken to complete all the tools were 45 minutes. They were informed for follow up after 3 months with the investigation reports. A file, Notepad, one pencil and one pen were given to each subject for participation and recording of consumption of medicines every day The subjects were informed over phone for follow up after three months at the OPDs. The tool II and Tool III were re-administered by interview technique and one or two visit by telephonic follow up at three (3) months, at six (6) months and at nine (9) months period. The data collection was completed after delivering thanks to the participant for their cooperation.

## Plan for data analysis

### The collected data would be analyzed by using the descriptive and Inferential Statistics after coding

ϖ Potential factors related, Socio-demographic data, clinical profile, therapy related, person related and health care delivery related data would be analyzed by descriptive statistics in the form of frequency and percentage, unpaired “t” test, Fisher’s exact test/ chi square test to check homogeneity between CABG and PTCA group,
ϖ Adherence to drug regimen related data would be analyzed by frequency, percentage, mean, and standard deviation. Cochran’s Q test for intergroup comparison and Chi square test for comparison between CABG and PTCA group,
ϖ Binary logistic regression analysis would be computed for prediction of relationship between potential factors with adherence to drug regimen
ϖ Quality of life related data would be transformed against the score of 100 for each item [Subtotal score would be computed under physical component summary(PCS) and mental component summary (MCS) score. Each domain of eight domains would be analyzed by descriptive statistics. Friedman’s Anova and Mann Whitney U test would be computed to find out significant difference in Quality of life with time interval between patient with CABG and PTCA.
ϖ Fisher’s exact test/ chi square test would be computed to find association between adherence to drug regimen and quality of life with selected demographic variables.
ϖ Coefficient of correlation would be computed to find relation of PCS and MCS of Quality of life score with adherence to drug regimen.

## Major findings of the study

### Demographic Characteristics

- The average age of the 355 patients who have undergone myocardial revascularization was 56.12±0.45 years
- Majority (84.51%) of the patients who have undergone myocardial revascularization were male.
- majority (90.70%) of the patients who have undergone myocardial revascularization were married.
- maximum (31.83%) and (33.52%) of the patients who have undergone myocardial revascularization had primary and secondary level of education respectively.
- Majority of patients (59.72 %) were full time worker.
- Majority (81.97%) of the patient belonged to single family.
- Majority (82.82%) of the patients who have undergone myocardial revascularization were living at rural areas
- Majority (80%) of the patients who have undergone myocardial revascularization had monthly family income below Rs. 20,000/-
- Majority (92.7%) of the patient were non-vegetarian.
- Majority (80.28%) of the patients who have undergone myocardial revascularization were smoker.
- Majority (73.80%) of the patients who have undergone myocardial revascularization had history of alcohol consumption.
- Majority (79.15%) of the patients who have undergone myocardial revascularization had no family history of coronary artery disease.
- Majority of the patient (85.07%)had normal BMI
- Maximum (45.92%) and (42.53%) of patient were in the group of NYHA classification II and I respectively.
- Maximum (78.87%) of the patient’s heart rate were within ≤10bpm increase from baseline.
- Majority (81.40%) of the patients’ SBP remained static.
- Majority (70.42%) of the patient’s LVEF was above 40%
- Majority (73.23%) of the patient’s one vessel was affected.
- Average duration of hospitalization was 9.6±1.53 days.
- All the patients had comorbid disease conditions as 80.56% had HTN, 12.11% had Type II DM,2.53% had Hypothyroidism, 0.84% had previous brain stroke and 3.385 patients had Arthritis and lung diseases.
- Average number of daily medication consumed was 6.61± 0.074 pcs
- Antiplatelet, antianginal, beta blocker and Antilipidemic drugs were prescribed for 100% patients with CABG and PTCA. ACE inhibitor group of drugs were prescribed for 24.02% and 58.52% patients with CABG and PTCA respectively, Calcium channel blockers group of drugs were prescribed for 28.49% and 85.22% of patients with CABG

and PTCA respectively. Diuretics were prescribed for 28.49% and 81.25%, Digoxin 0.01% by both the groups, antiarrhythmic group of drugs by 0.05% and 11.36% patients with CABG and PTCA . The other group of medications (pan 40, Antibiotic Mupirocin ointment, Tab paracetamol Syr. lactose, Tab Alprazolam, Tab. Thyronorm) (any one) were prescribed for 100% patients with CABG and PTCA.

- Majority (70.39% and 59.65%) patients with CABG and PTCA respectively were taking medicine with the help of other members like wife, son, daughter, daughter in law and son-in law)
- The average number of medication unavailable from the hospital medicine counter was 3.40 ± 0.06 for patient with CABG and 2.85 ±0.05 for patients with PTCA
- The average Cost of purchasing unavailable medication from Govt. medicine counter was Rs.819.55±251.97for patients with CABG and Rs. 818.18±18.98 for patients with PTCA.
- The average knowledge about drugs for both the group of patients with CABG and PTCA were 4.715±0.05 and 4.710±0.05 respectively.
- The Information provided by the medicine counter regarding time of taking medicines was 100% as responded by both the group of patients with CABG and PTCA. But only 0.03% and 0.01% patients with CABG and PTCA received information regarding}effects of medicine
- The patient who have undergone CABG or PTCA were homogeneous in respect of age, marital status, educational status, employment status, Total monthly family income, history of smoking and history of alcohol consumption, BMI, heart rate/pulse, LVEF, findings of chest auscultation, number of vessels affected, no of medication consumed, person administering medication, Approximate monthly cost of purchasing unavailable medicine from Govt. medicine counter, Patients knowledge about drugs, Information provided by the medicine counte**r.**
- The patient who have undergone CABG or PTCA were heterogeneous in respect of gender, type of family, place of residence and family history of heart disease, NYHA classification, Systolic Blood pressure, duration of hospitalization, number of comorbid disease condition and number of medication unavailable from the hospital medicine counter

## Adherence to drug regimen

- Majority (82%) of the patients with CABG and PTCA were adherent to drug regimen at initial month after revascularisation. The percentage of adherence with medication was74% after 3 months, 75% after 6 months, and 59 % after 9 months among patients with CABG whereas percentage of adherence among patient with PTCA was 66% after 3 months, 63% after 6 months and 44 % after 9 months
- The average score of adherence to drug regimen was 12.24±.03 in initial month, 12.77±0.08 after 3 months, 12.8±0.08 after 6 months and 15.47±0.02 after 9 months among total patients who have undergone myocardial revascularisation either by CABG or PTCA.
- The calculated “t” test and “F” test result showed that there is significant difference found between level of adherence to drug regimen among patient with CABG and PTCA after 3 months, 6 months and 9 months’ interval. (p<0.001). [**refer table no 1**]

### Association between adherence to drug regimen with potential factors

- The level of adherence to drug regimen was independent and not associated with age, age, gender, marital status, educational status, employment status, total monthly family income, place of residence, type of family, history of smoking, history of alcohol consumption, family history of heart disease with BMI, NYHA classification, heart rate, systolic BP, LVEF, Number of vessels affected, duration of hospitalization, number of comorbid disease conditions, person administering medication, Information provided by the medicine counter regarding effects of medicin**es,** number of medicine consumed per day and cost of medicine to buy unavailable medicine from outside.
- Number of drugs unavailable in Govt. medicine counter was associated with adherence to drug regimen among patients with myocardial revascularisation.
- Age (OR= 0.9405, p=0.0263) and educational status (OR= 0.7061 p=0.0343) were significant predictors of adherence to drug regimen among patients with PTCA. [**refer table no 2]**

### Quality of life of patients who have undergone Myocardial revascularisation

- The Physical component score of quality of life was improved from initial month to nine months among patients who have undergone CABG and PTCA. (p<0.001) [**refer table no 3**]
- The Mental Component score of quality of life was improved from initial month to nine months among patients who have undergone either CABG or PTCA. (p<0.001) [**refer table no 4**]
- The calculated Chi square / Fisher exact test result revealed that Physical component summary and Mental component summary of quality of life were not associated with age, gender, marital status, educational status, employment status, total monthly family income, place of residence, type of family, history of smoking, history of alcohol consumption and family history of heart disease

### Relationship of adherence to drug regimen with Quality of Life

- The calculated correlation of coefficient result showed that PCS of quality of life (p=.007, p=0.002, p=0.04) and also at 6 months’ time MCS, of Quality of life were negatively related with adherence to drug regimen. (p<0.008) [**refer table no 5**]

## Discussion

The findings of the present study were discussed based on the objectives and hypotheses of the study and compared with the findings of similar and contradictory studies.

Out of 355 patients with myocardial revascularization, the average age of the 179 patients who have undergone CABG was56.25±0.59, majority (90.50%) were male and (94.97%)were married and 176 patients who have undergone PTCA was 56.25±0.69 years where majority (78.41%) were male and (90.70%) married

The findings of the present study were in accordance to Loponen P, Luther M, Korpilahti K, Wistbacka JO, Huhtala H, Laurikka J, et al. who conducted the research study to find “HRQoL after coronary artery bypass grafting and percutaneous coronary intervention for stable angina”. The average age of the patients with CABG was 66.1 years and patients with PTCA was 64.5years, majority were male (79.5%) and married (68.6%).

Aonther study conducted by Allahbakhshian A, Gharamaleki RN, Rahmani A, Tabrizi FJ, Allahbakhshian M, Gholizadeh L. to find “Is Beliefs About Medication a Factor in Adherence to the Medicine in Patients Undergoing Coronary Angioplasty” and the result showed that average age of the patient was 56.99±12.88 years,78.7% were male and 89.3% were married. **T**he similarity of findings revealed in the research studies conducted by . Pacaric S, Turk T, Eric I, Orkic Z, Eric AP, Milostic-Srb A. et al.[36], Salari A Balasi LR, Ashouri A, Moaddab F, Zaersabet F, Nourisaeed A. [33] Yan BP, Chan LLY, Lee VWY, Yu M, Wong MCS, Sanderson J. et al[51]

The findings of the present study were contradicted with the findings of a research study conducted by Colosimo CF, De Sousa AG, Da Silva GS, Piotto RF, Pierin AMG. Arterial hypertension and associated factors in patients submitted to myocardial revascularization at Brazil, as average age of the patients undergoing Myocardial revascularisation was 62.43±9.47 years. [52]

The present study also showed that the maximum (31.83%) and (33.52%) had primary and secondary level of education respectively, along with that majority (82.82%) of the patients were living at rural areas. The similar findings were seen in several research studies conducted in India [11,12,25]

The data related to employment status shows that at the time of admission maximum (45.25%) patients with CABG were full time worker,15.08%were part time worker, 39.67% were disable/ not working due to disease condition or retired from the job. But majority (74.43%) patients with PTCA were full time worker at the time of admission,5.68% were part time worker,19.89%were disabled /not working due to disease condition or already retired and (80%) had monthly family income below Rs. 20,000/- and (92.7%) of the patient were non-vegetarian.

Majority (80.28%) of the patients who have undergone myocardial revascularization were smoker and (73.80%) had history of alcohol consumption, majority (79.15%) of the patients who have undergone myocardial revascularization had no family history of coronary artery disease which is similar with the findings of several studies conducted in India [12]

The data related to clinical variables shows that majority of the patient (85.07%)had normal BMI, maximum (45.92%) and (42.53%) of patient were in the group of NYHA classification II and I respectively, maximum (78.87%) of the patient’s heart rate were within ≤10bpm increase from baseline, majority (81.40%) of the patients’ SBP remained static and (70.42%) of the patient’s LVEF was above 40%, majority (73.23%) of the patient’s one vessel or one coronary artery was affected.

The admission data also revealed that average duration of hospitalization was 9.6±1.53 days.

All the patients had comorbid disease conditions as 80.56% had HTN, 12.11% had Type II DM,2.53% had Hypothyroidism, 0.84% had previous brain stroke and 3.38% patients had Arthritis and lung diseases.

The number of medicine consumption related data showed that average number of daily oral medication consumed by the patient who have undergone myocardial revascularisation was 6.61± 0.074 pcs. The similar findings also observed in maximum number of studies conducted to find level of adherence to drug regimen. [31,34,53,54]

### The first objective of the study was to assess adherence to drug regimen among patient who have undergone myocardial revascularization

The average score of adherence to drug regimen was 12.24±.03 in initial month, 12.77±0.08 after 3 months, 12.8±0.08 after 6 months and 15.47±0.02 after 9 months among total patients who have undergone myocardial revascularisation either by CABG or PTCA.

The subgroup analysis shows that majority (82%) of the patients with CABG and PTCA were adherent to drug regimen at initial visit but reduced with time. The percentage of adherence with medication was74% after 3 months, 75% after 6 months, and 59 % after 9 months among patients with CABG whereas percentage of adherence among patient with PTCA was 66% after 3 months, 63% after 6 months and 44 % after 9 months.

The calculated “p “value shows that significant difference in level of adherence exists between 1 month, 3 months, 6 months and 9 months’ time period among total group of patients undergone myocardial revascularisation and also with individual group of patients with CABG or PTCA. So the level of adherence to drug regimen was decreased with time interval.

### The findings of the present study were supported by the findings

The similar monocetric prospective observational study was conducted by Pietrzykowski L, Michalski P, Kosobucka A Kasprzak M, Fabiszak T, Stolarek W, et al. on medication adherence and its determinants in patients after myocardial infarction treated with primary coronary intervention(PCI) The result revealed that during 1-year follow-up, adherence for all three drug classes was 64 ± 25%, with 67 ± 32% for ACEIs, 62 ± 34% for P2Y12 receptor inhibitor and 64 ± 32% for statins. A gradual decline in adherence was observed from 65% ± 26% in the first quarter of follow-up to 51% ± 34% in the last quarter of follow-up (p < 0.00001). Sufficient adherence for all drugs classes was found only in 29% of patients throughout the whole follow-up period (44% for ACEI, 36% for P2Y12 receptor inhibitor and 41% for statins).

A similar study conducted by Salari A Balasi LR, Ashouri A, Moaddab F, Zaersabet F, Nourisaeed A. on Medication Adherence and its Related Factors in Patients Undergoing Coronary Artery Angioplasty showed that 75 patients (28%) did not adhere to their medication [33]

Another similar study was conducted by Gonarkar SB, Dhande P on Medication adherence and its determinants in myocardial infarction patients: An Indian scenario. The result shows that medication adherence was significantly reduced from 87.1% at 1st month to 57.4% at 6th month post-MI. After applying McNemar test, this reduction in good adherence in the study subjects over the 6-month period was found to be statistically significant (*P* < 0.0001).[1]

### The second objective was to find potential factors of adherence to drug regimen among patients who have undergone myocardial Revascularization

The findings of the present study revealed that Age (OR= 0.9405, p=0.0263) and Educational status (B coefficient -0.34795 OR= 0.7061 p=**0.0343**) were significant predictors of adherence to drug regimen among patients with PTCA. The result showed that increasing age lead to increasing adherence to drug regimen, may be with experience by understanding the benefit of medicines. The findings also revealed that better adherence to drug regimen in patient with education below primary level. This might be due to their confidence on Physicians’ advice and fear of complications and associated extra financial burden on their family.

A similar study conducted by Pietrzykowski L, Michalski P, Kosobucka A Kasprzak M, Fabiszak T, Stolarek W, et al. on medication adherence and its determinants in patients after myocardial infarction treated with primary coronary intervention(PCI)revealed that in multivariate analysis, age, prior CABG, level of education, place of residence, economic status and marital status were independent predictors of drug adherence. Whereas patients > 65 years and having a history of prior CABG more often had an insufficient adherence to drugs, married and hypertensive patients, city inhabitants and patients with higher education tended to have a sufficient drug adherence. A number of socioeconomic and clinical factors have been identified to affect medication adherence over time. [34]

Another study conducted by Aghabekyan S, ThompsonME, Arahamyan L. on medication noncompliance and patient satisfaction following percutaneous coronary intervention showed that age, gender, health status, smoking status, and cost were associated with medication noncompliance (P < 0.05). Medication noncompliance was positively related to cost (odds ratio [OR]= 2.57, 95% CI = 1.33-4.97) and inversely related to health status (OR = 0.46, 95% CI = 0.25-0.85) and age (OR = 0.94, 95% CI = 0.91-0.97).[]

The similar findings was also observed in the study conducted by Balasi RL, Paryad E, Booraki SH,Leili EK, Meibodi AMS, Sheikhani NN. Medication adherence after CABG and its related to medical belief with the aim of the study to determine relationship medication belief and adherence 6 months after CABG. The result showed a significant relationship between adherence to medication with age(p<0.001), marital status(p<0.02),educational attainment<0.00001) family history of heart disease and residence area. [24]

### The third objective was to assess the quality of life among patients who have undergone myocardial revascularization

The Physical component summary (PCS) and Mental Component summary (MCS) scores are two meta-scores of SF-36 calculated from the SF-36 questionnaire and reflect a patient’s overall physical and mental health status. PCS is consisted of five domains in questionnaire as physical functioning, role physical, bodily pain, vitality and general health . MCS is consisted of three domains as role emotional, social functioning and mental health. The mean and standard deviation (62.42±12.74) in general health was maximum in 9 months than in one months(36.20±9.75), three months(51.28±13.14) and six months(57.10±14.74) period. In physical functioning domain, The mean and standard deviation (60.99±11.74) in general health was maximum in 9 months than in one months(29.15±14.96), three months(55.01±13.07) and six months(60.14±11.74) period. In role physical functioning physical, the mean and standard deviation (34.93±26.67) was maximum in 9 months than in one months(6.20±17.54), three months(10.07±20.56) and six months(34.30±25.93) period. In social functioning, the mean and standard deviation (66.94±16.59) was maximum in 9 months than in one months(36.55±18.68), three months(57.29±23.60) and six months(65.81±18.01) period. In bodily pain, the mean and standard deviation (70.35±13.79) was maximum in 9 months than in one month (44.54±19.61), three months(65.13±18.89) and six months(68.91±15.64) period. . In energy, the mean and standard deviation (68.59±10.86) was maximum in 9 months than in one months(58.19±12.88), three months(62.44±12.32) and six months(66.83±10.84) period. . In emotional wellbeing, the mean and standard deviation (63.40±9.24) was maximum in 9 months than in one months(49.75±10.79), three months(59.36±11.05) and six months(61.48±10.69) period.

The PCS and MCS score of patient with myocardial revascularisation was improved from within one month (116.09±37.44, 149.74±33.35) to 9 months (228.69±37.16, 232.17±43.16) after intervention or surgery of myocardial revascularisation. The p value shows significant difference in 4 time intervals of 1 month, 3 months, 6 months and 9months in all eight domains and also in PCS and MCS among patient who have undergone CABG or PTCA.

The similar findings were observed in the study conducted by Ambina K, Shalimol US, Anjana AP . on Quality of life(QOL) among post CABG patients coming for follow up at CTVS OPDS of Amrita Institute of Medical Science, Kochi. The study findings showed that 97% had good quality of life in PCS domain and 68.3% in the domain of MCS. [37]

The similar findings were also observed in the study conducted by Peric V, Stolic R, Jovanovic A,Grbic R, Lazic B, Sovtic S et al.Predictors of quality of life improvement after 2years of Coronary Artery Bypass surgery .The result showed the improvement in all sections of QOL two years after CABG [38]

### The fourth objective was to find association between adherence to drug regimen with selected demographic variables among patients who have undergone myocardial Revascularization

There was no association between adherence to drug regimen with selected demographic, variables, condition related, therapy related variables, patient related and provider and health care delivery system related variables except number of drugs unavailable in Govt. medicine counter, which was associated with adherence to drug regimen among patients with myocardial revascularisation. It showed that patients are dependent on Government hospital medicine supply as cost of purchasing of unavailable medicines from Govt. medicine counter remains unaffordable for them.

The findings of the present study was supported by the study conducted by Dusetzina SB, Besaw RJ, Whitmore CC, et al. Cost-Related Medication Nonadherence and Desire for Medication Cost Information Among Adults Aged 65 Years and Older in the US in 2022. The study result revealed that cost-related medication nonadherence was 20.2%. Some participants used extreme forms of cost-coping by foregoing basic needs (8.5%) or going into debt (4.8%) to buy or afford medications. 80% of respondents with cost-related nonadherence reported that If the actual price was much more than the estimated real-time benefit tool price, nearly it would affect their decision to start or keep taking a medication. [62]

The finding of the present study was also supported by the study of Pandey KR, Meltzer DO. On financial burden and impoverishment due to cardiovascular medications in low and middle income countries: An illustration from India revealed that the more expensive cardiovascular medicine regimens could be unaffordable to as much as 81% of the rural and 58% of the urban population in India if they wanted to purchase these medicines out of pocket. If all of the 93 million of the rural and 76 million of the urban adult population who may benefit from cardiovascular medicines were to buy these medications out of pocket, as many as 45 million rural and 30 million urban Indians could be financially burdened with impoverishment of 17 million rural and 10 million urban people. These figures represent an increase in baseline poverty levels by 2 percentage points for rural India and 3.2 percentage points for urban India. Poverty gaps could increase by 2.91 percentage points in rural India (from a baseline of about 7.6%) and 2.88 percentage points in urban India (from a baseline of about 6.2%). As a percentage of baseline poverty levels, the poverty ratios could increase by about 6% and 11% for rural and urban areas respectively. However the poverty gap indices could increase by about 38% of baseline for rural India and by about 46% for urban India. This shows that the financial burden posed by cardiovascular medication purchase could be significant and much more severe than that represented by the increase in the number of people pushed below the poverty line alone. If 75% of these people were to buy cardiovascular medicines out of pocket, as is the current share of out of pocket health expenditures in India, as many as 34 million rural Indians and 22 million urban Indians could find these medicines unaffordable with resulting impoverishment of 13 million rural Indians and 7 million urban Indians.

### The fifth objective was to find association between quality of life with selected demographic variables among patients who have undergone myocardial Revascularization

There was no association between adherence to drug regimen with selected demographic, variables.

There was improvement in Quality of life due to surgical or interventional procedure, which might be reason for not relating with socio-demographic variables.

### The sixth objective was to find correlation between adherence to drug regimen with quality of life among patients who have undergone myocardial Revascularization

The calculated correlation of coefficient result showed negative relation with a very weak magnitude between PCS of quality of life (p=.007, p=0.002, p=0.04) and also at 6 months’ time MCS, of Quality of life with adherence to drug regimen. (p<0.008). This might be due to the different perception about the disease, its treatment and prognosis. The graft or stent had enhanced the blood flow on the affected coronary artery, Consequently, the absence of symptoms of CAD had been perceived as medications are not necessary, it is the sign of healing, so the patient had a good quality of life but low adherence to medication.

A similar retrospective cohort study conducted by Hasandokht T, Salari A. on Medication adherence and quality of life in coronary artery bypass grafting patients revealed that Physical and mental components of QOL were negatively associated with medication (B:-0.18, p:0.04; B:-0.29, p:0.02, respectively) and follow-up visit observance (B:-0.3, p:0.01; B:-0.3, p: 0.01, respectively). [19]

### The present study was contradicted by

A cross sectional study conducted by Coelho M, Costa ECA, Richter VC, Ciol MA, Schmidt A, Dantas RAS. et al. Perceived health status and pharmacological adherence of patients who underwent percutaneous coronary intervention. The result revealed positive correlation of moderate magnitude between measurements of pharmacological adherence and perceived health status.[31]

### Other findings

The fundamental reasons for nonadherent to cardiovascular medication varied among patients who have undergone Myocardial revascularisation. Though it was not assessed by structured tools but during interview it was revealed that inadequate health literacy regarding cardiovascular surgery or Interventions as it was mistakenly believed by the patient as getting cured, ineffective working relationship with pharmacist of medicine counter with the patient raises demands for discussion with patient regarding drug administration schedule, intended benefits, adverse reactions and cost to buy unavailable medicines, the conditions like cognitive, vision and hearing impairment may dispose the patient with older age in non-adherence group, the cost to buy unavailable medicines imposes burden on the family for refilling medicines and inefficacy of medicines supplied, side effects of medication, depression, anxiety related to ineffective recovery to join in previous occupation also need to be addressed for better adherence.

### Conclusion

Based on the findings, the present study concluded that level of adherence to drug regimen decreases with time interval from within one month to 9 months for all the patients undergoing myocardial revascularisation either in the form of CABG or PTCA.

The adherence to drug regimen was associated with no of drugs unavailable from Govt. Medicine counter.

The level of adherence to drug regimen was independent and not associated with socio demographic, clinical person related and health care delivery system related factor except Age and educational status were significant predictors of adherence to drug regimen among patients with PTCA.

The quality of life of patients who have undergone myocardial revascularisation improved with time interval from within one (1) month to nine (9) months for all the patients who have undergone myocardial revascularisation and also significantly different between patients with CABG and PTCA.

The physical component of quality of life was negatively associated with medication adherence from 3 months to 9 months and the mental component part of quality of life was associated with adherence to drug regimen at six (6) months.

## Limitation

The findings of the present study could not be generalized due to the following reasons

- The study was conducted in a single setting and only from the middle and lower income group of patients who were attending cardiac OPD of Government hospital, which may not be generalizable to the higher income group and receiving follow up services from private or superspeciality hospitals.
- Adherence to drug regimen was measured through self-reporting, which may be liable for recall biasness and social desirability response biasness.
- Sample was selected by non-probability convenience sampling technique

## Recommendation

The following recommendations were made based on the present study findings

- A similar study can be conducted by taking sample from different tertiary care hospitals.
- A comparative study can be conducted to assess the adherence to drug regimen among patients attending cardiac Outpatient Department of Government and Private hospitals.
- A study can be conducted to find effectiveness of different electronic devices for improvement in medication adherence among patients who have undergone myocardial revascularisation.
- A study can be conducted to find effectiveness of a learning package on health literacy in terms of knowledge and practice for increased medication adherence among patient who have undergone myocardial revascularisation.
- A cross sectional survey can be conducted to find relation between level of medication adherence and quality of life in a large scale among low literacy group.
- A similar study can be conducted to find predictors of Quality of life among patients who have undergone myocardial revascularisation.
- Values are presented as Mean±SD for numerical variables and as count (%)for categorical variables
- P value of last column denotes intergroup comparison by Cochran’s Q test and last row denotes between group comparison by chi square (ꭕ^2^) test as appropriate

**Table 1.**
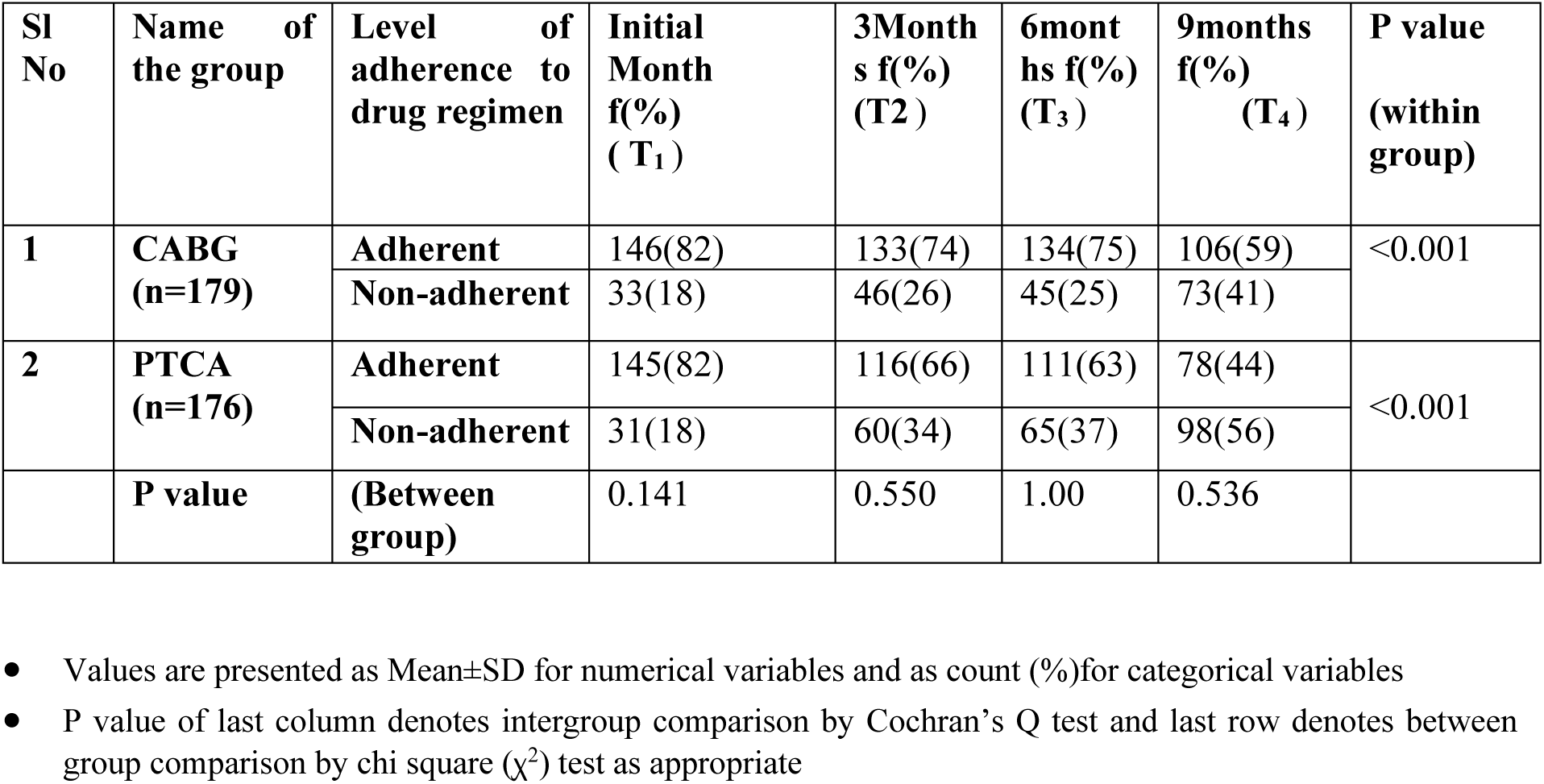
Frequency and percentage distribution of subjects based on level of adherence to drug regimen n =355.

**Table 2.** Binary logistic regression analysis for association between level of adherence with related factors among patients with PTCA n=176.

| Sl no. | Types of variables | Coefficient | Std. Error | Wald | P | Odds ratio | 95% CI |
| --- | --- | --- | --- | --- | --- | --- | --- |
| **Demographic Variables** |  |  |  |  |  |  |  |
| 1 | Age | -0.06131 | 0.02759 | 4.9355 | **0.0263** | 0.9405 | 0.8910 to 0.992 |
| 2 | Gender | -0.10675 | 0.63993 | 0.02783 | 0.8675 | 0.8987 | 0.2564 to 3.150 |
| 3 | Educational Status | -0.34795 | 0.16438 | 4.4806 | **0.0343** | 0.7061 | 0.5116 to 0.974 |
| 4 | Employment | 0.46238 | 0.31884 | 2.1030 | 0.1470 | 1.5879 | 0.8500 to 2.966 |
|  | status |  |  |  |  |  |  |
| 5 | Monthly_family Income | -0.41172 | 0.27195 | 2.2921 | 0.1300 | 0.6625 | 0.3888 to 1.129 |
| **Clinical variables** |  |  |  |  |  |  |  |
| 6 | LVEF % | 0.6573 | 0.41037 | 2.5655 | 0.109 | 1.9296 | 0.8633 to 4.313 |
| 7 | Chest_Auscultation | 0.3827 | 0.63168 | 0.3670 | 0.544 | 1.4662 | 0.4251 to 5.057 |
| 8 | Duration_of_Hospitalization | -0.0390 | 0.094100 | 0.1719 | 0.678 | 0.9617 | 0.7998 to 1.156 |
| 9 | No_of_Comorbidities | -0.3046 | 0.28322 | 1.1567 | 0.282 | 0.7374 | 0.4233 to 1.284 |
|  | Constant | 0.7921 | 1.46153 | 0.2938 | 0.587 |  |  |
| **Medicine related, Person related and healthcare delivery system related variables** |  |  |  |  |  |  |  |
| 10 | Person giving medications | 0.0402 | 0.15824 | 0.06467 | 0.79 | 1.0411 | 0.7634 to 1.419 |
| 11 | No of daily drugs | 0.1409 | 0.18016 | 0.6119 | 0.43 | 1.1513 | 0.8088 to 1.638 |
| 12 | Number of drugs unavailable in Govt. medicine counter | 0.6851 | 0.40816 | 2.8178 | 0.09 | 1.9841 | 0.8915 to 4.415 |
| 13 | Knowledge related to medication | 0.5672 | 0.75211 | 0.5688 | 0.45 | 1.7634 | 0.4038 to 7.701 |
| 14 | Cost to buy unavailable medicines | -0.0003 | 0.001337 | 0.05312 | 0.82 | 0.9997 | 0.9971 to 1.002 |
|  | Constant | 1.8806 | 1.60403 | 1.3747 | 0.24 |  |  |

**Table 3.** Comparison of physical component summary (PCS) and Mental Component summary(MCS) of quality of life between patients with CABG and PTCA n=179+176.

| Sl No | Name of the group | Physical component summary(PCS) |  |  |  | “p” value |
| --- | --- | --- | --- | --- | --- | --- |
|  |  | 1 month | 3 months | 6 months | 9 months |  |
| 1 | CABG(N=179) | 130.3±32.07 | 208.7±28.14 | 239.7±30.70 | 244.9±30.55 | <0.001 |
| 2 | PTCA(n=176) | 101.6±37.13 | 153.86±30.40 | 200.9±36.97 | 212.2±36.23 | <0.001 |
|  | P value | <0.001 | <0.001 | <0.001 | <0.001 |  |

- Friedman test for within group and Mann Whitney U test for between groups.

**Table 4.** Comparison of Mental Component summary(MCS) of quality of life between patients with CABG and PTCA n=179+176.

| Sl No | Name of the group | Mental Component summary(MCS) |  |  |  | “p” value |
| --- | --- | --- | --- | --- | --- | --- |
|  |  | 1 month | 3 months | 6 months | 9 months |  |
| 1. | CABG(N=179) | 156.6±32.53 | 209.7±30.17 | 249.4±36.40 | 252.6±34.83 | <0.001 |
| 2. | PTCA(n=176) | 142.8± 32.92 | 162.7±32.44 | 190.4±32.65 | 211.3±40. | <0.001 |
|  |  | <0.001 | <0.001 | <0.001 | <0.001 |  |

- Friedman Anova for within group and Mann Whitney U test for between groups.

**Table 5.** Correlation matrix between PCS and MCS of Quality of Life with adherence to drug regimen in four time points with scatter diagrams n=355.

| Sl No | Correlational matrix (Spearman Rho and p value) | Within 1 month | 3months | 6 months | 9 months |
| --- | --- | --- | --- | --- | --- |
| 1 | Adherence to drug regimen score with PCS | 0.07367<br>P=0.1660 | -0.1411<br>P=0.0077 | -0.1934<br>P=0.0002 | -0.1088<br>P=0.0405 |
| 2 | Adherence to drug regimen score with MCS | 0.1006<br>P=0.0582 | -0.09843<br>P=0.0639 | -0.1393<br>P=0.0086 | -0.07468<br>P=0.1603 |

## Data Availability

ADHERENCE TO DRUG REGIMEN AND QUALITY OF LIFE

## Funding

Self –Financed. No fund was allocated for this study

## Conflict of interest

There is no conflict of interest.

## Notes

### Competing Interest Statement

The authors have declared no competing interest.

### Clinical Trial

NOT APPLICABLE OBSERVATIONAL STUDY

### Funding Statement

SELF FINANCED NO FUND FROM ANY SOURCE.

### Author Declarations

Institutional Ethics Committee of NRS Medical College and Hospital

